# Global Disparities in Access to Dermatological Care: the Skin Health Observatory

**DOI:** 10.64898/2026.02.06.26345759

**Authors:** Esther E. Freeman, Joseph M. Yardman-Frank, Jennifer Kilmer, Ann Pacheco, Karry Su, Devon E. McMahon, Christine Li, Sarah Anwar, Kathryn Barger, Yiqi Qian, Alexis Strahan, Simon Westby, Ramesh Bhat, Mahira El Sayed, Wendemagegn Enbiale, Cristina Galvan-Casas, Xinghua Gao, Srie P. Gondokaryono, Abdul G. Kibbi, Adriene Lee, Fatimata Ly, Jorge Ocampo-Candiani, Marie-Aleth Richard, Ricardo Romiti, Henry W. Lim, Junko Takeshita, Delphine Kerob, Bertrand Chuberre, Guénolée de Lambert, Lucinda C. Fuller, Christopher E.M. Griffiths, Ncoza C. Dlova

## Abstract

**Background:** Skin disease affects 4.7-4.9 billion individuals globally; however, little is known about access to dermatological care.

**Methods:** We conducted a multinational, cross-sectional survey of dermatological care across 194 WHO member states and three additional geographic areas in 2024-2025. Primary outcomes included dermatologist density per 100,000 population and number of dermatologists globally. Secondary outcomes included training programme density, workforce distribution, perceived access to care, and health system characteristics. Descriptive statistics and nonparametric tests compared outcomes across World Bank Income (WBI) levels and WHO regions.

**Findings:** Responses were obtained from 158 countries. Mean dermatologist density was 2.66 per 100,000, ranging from 0.37 in low-income (LICs) to 5.05 in high-income countries (HICs). There are estimated 175,633 dermatologists globally (95% CI: 173,598-177,668). Forty-two percent of countries reported inadequate or extremely poor access to dermatological care. There was significant variation (p < 0.001) in access to all types of subspecialty care (paediatric, surgical, dermatopathology) across WBI levels, with consistently worse access in lower-income countries. Dermatologists are primarily based in urban centres (79%). Twenty-one percent of countries lack dermatology training programs, with training varying by WBI level (p < 0.001). Non-dermatologist healthcare workers bear a substantial responsibility for management of skin disease.

**Interpretation:** Significant global disparities exist in access to dermatological care, particularly in lower resource settings. Achieving skin health equity will require global commitment to expanding/funding training programmes, incentivizing decentralization of dermatology practice, and optimizing alternative care delivery including upskilling front-line healthcare workers.

**Funding:** International League of Dermatological Societies and L’Oreal Dermatological Beauty.

## Introduction

Skin diseases constitute one of the most pervasive yet overlooked drivers of global morbidity. There are approximately 4.7-4.9 billion people living with skin disease, accounting for over 42.9 million disability-adjusted life-years.^1, 2^ The burden of skin disease disproportionately affects children and individuals in low- and middle-income countries (LMICs).^2^ The visibility of dermatological conditions contributes to patient stigma, resulting in mental health comorbidities, discrimination, and reduced educational and economic opportunities.^3, 4^ Despite being a leading cause of disability worldwide, dermatological disease has historically received limited attention in global health policy.^5-7^

In May 2025, the World Health Assembly adopted a resolution on ‘Skin diseases as a global public health priority.’^1^ The resolution frames skin health as essential to universal health coverage (UHC), acknowledges the substantial burden of skin disease, and calls for additional financing, surveillance, and workforce capacity.^1^ Building on this momentum, the *Lancet* Commission on Skin Health was established in November 2025, with the aim of forming a framework and action plan for global skin health governance.^9^ Achieving these goals hinges on understanding the state of access to dermatological care worldwide and accurate assessment of the dermatology workforce.

Access to dermatological care worldwide and the capacity to provide essential dermatology services have not been systematically studied. Prior limited regional studies reveal striking disparities in dermatologist and training programme distribution, a marked urban-rural misdistribution, and a disproportionate number of dermatologists in the private sector. These disparities affect both high-income countries (HICs) and low-income countries (LICs).^10, 17, 18^

To address the critical gap in global dermatology workforce data, the International League of Dermatological Societies (ILDS) designed the Global Access to Skin Health Observatory (SkinObservatory) study, which represents the first systematic global assessment of access to dermatological care across World Health Organization (WHO) member states. The SkinObservatory study aims to evaluate the capacity, structure, and barriers to dermatological care by assessing workforce shortages, training capacity, and infrastructure.

In this report, we describe global, regional, and income-group patterns in dermatologist density and training capacity; examine imbalances in dermatologist distribution among urban vs rural areas and the public vs private sectors; characterize access to, and primary providers of, general and specialized dermatological care. These findings will inform strategies to strengthen dermatology workforce development and guide national and regional planning to advance equitable access to skin health services.

## Methods

### Study design

We conducted a multinational, cross-sectional survey study across all 194 WHO member countries, the occupied Palestine territory (recognised by the WHO but not a full member)^21^, Chinese Taipei (Taiwan) and Hong Kong, China (both captured under the WHO member state China but with independent dermatological societies). Herein, we use the term ‘country’ to refer to these 197 separate geographic units.

We devised a multicomponent leadership structure composed of a steering committee, regional directors, and a technical advisory group. The steering committee comprised seven global health dermatology leaders from five countries, including senior-level leaders from the WHO and ILDS, two core study design researchers, and an epidemiological methods director. The regional directors consisted of 13 senior global health dermatology leaders, with representation from each WHO region.

The study was reviewed and deemed exempt by the Massachusetts General Brigham Institutional Review Board (Protocol #2024P001321).

### Study population

The study survey was distributed to key dermatology stakeholders in each country; for countries with ILDS member societies, it was delivered to national society leaders. For countries without national societies, dermatology leaders were identified through our global network of collaborators and the WHO. In countries with no dermatologists, the survey was distributed to WHO country programme officers.

### Procedures

Initial survey content was developed through literature review and global stakeholder meetings. The survey was based on the WHO’s health systems ‘Building Blocks’ framework^22^ and informed by the Global Kidney Atlas as a tool to assess access to specialty care (Supplement 1.1).^23-25^

Next, a two-round modified Delphi process was conducted with 20 panelists from the steering committee and regional directors to reach consensus on draft questions. Questions reaching 75% agreement were included in the revised survey;^26^ those that did not were removed or amended based on panellists’ comments. This yielded the official 47-question SkinObservatory survey, which was available in English, French, Spanish, and Portuguese.

The survey was distributed online via REDCap through the ILDS, with data collection and follow-up conducted between August 2024 and October 2025. Study staff coordinated follow-up emails with non-responders at 3-month intervals.

Data were examined for incongruencies and outliers. Respondents were contacted for clarification as needed. For countries with multiple respondents, see Supplement 1.2 for the adjudication process. Following the initial round of data cleaning, all data were reviewed for validity and accuracy by the study team. Any remaining discrepancies were adjudicated by the study’s principal investigator and methods director.

Total population count and World Bank Income (WBI) level classification were extracted from the 2024 World Bank estimates (Supplement 1.3).^27, 28^ WHO region classification was derived from the official 2024 WHO Member States list.^29^

Reporting followed the WHO Guidelines for Accurate and Transparent Health Estimates Reporting (GATHER) statement.^30^ Data output will be available on www.skinobservatory.org.

### Outcome measures

The co-primary outcome measures were: (1) the density of dermatologists per 100,000 population per country; and (2) the estimated total number of dermatologists worldwide.

Secondary outcome measures included: the density of dermatology training programmes per 100,000 population; distribution of dermatologists by practice location and practice setting; perceived quality of access to dermatological care; distribution of different providers responsible for diagnosis and treatment of skin disease; national requirements to become a dermatologist; and distribution of perceived dermatological care provider shortage. Dermatological care was investigated across five different care domains – general, complex, surgical, paediatric and dermatopathological.

We also assessed additional workforce and health system characteristics, including care settings and locations, cosmetic dermatology, and government prioritisation of dermatological care.

### Statistical analysis

Descriptive statistics were calculated and reported as means or frequencies, as appropriate. Statistical testing of variation across WBI levels and WHO regions was performed using chi-square and Fisher’s exact tests for categorical variables and Kruskal-Wallis test for continuous and ordinal variables.

Each country’s density of dermatologists and dermatology training programmes were calculated per 100,000 population. The global density of dermatologists was calculated based on mean country density. To estimate the number of dermatologists worldwide, we predicted the number of dermatologists for countries that did not report a total using an interactive two-way ANOVA, with WBI level and WHO region as factors (Supplement 1.4).

To explore what constitutes a sufficient dermatologist density per country, we calculated the mean density of dermatologists among countries that reported having at least ‘Adequate’ access to comprehensive dermatological care. Using this density as a threshold, countries were labelled either ‘Sufficient’ or ‘Insufficient’. WBI levels were then collapsed into two groups: ‘high-income’ (all HICs) and ‘non-high-income’ (all other countries). A logistic regression was used to calculate the odds of having a sufficient density of dermatologists by income group.

Statistical analysis was carried out in STATA/SE (version 18.0), R (version 4) and SAS (version 9.4). Figures were produced by Graphicacy in collaboration with the study team.

## Results

The survey was completed for 158 countries, representing 96.7% of the global population and a cumulative response rate of 80.2% (Supplement 2.1 and 2.2).

### Distribution of countries

Responses included representation across all four WBI levels and six WHO regions. At each WBI level, a minimum of 76% of countries responded. Twenty-one countries were from the low WBI level, 45 from lower-middle income, 40 from upper-middle income, and 52 from HICs (13%, 29%, 25% and 33% of respondents respectively).

Across WHO regions, 40 responding countries were from the African region (AFR), representing 25% of respondents and 85% of African countries. Similarly, 19 responding countries were from the Eastern Mediterranean region (EMR) (12% of respondents, 86% of the region), 37 from the European region (EUR) (23% of respondents, 70% of the region), 22 from the Western Pacific region (WPR) (14% of respondents, 76% of the region), ten from the South-East Asian region (SEAR) (6% of respondents, 91% from the region), and 30 from the Americas region (AMR) (19% of respondents, 86% of the region).

### Density, demographics, and distribution of dermatologists

The country-level density of dermatologists ranged from 0 to 13.96 per 100,000 people. Nine countries reported having no dermatologists while Greece reported the highest density. Across responding countries, the mean density was 2.66 dermatologists per 100,000 people, but there was significant variation (p < 0.001) across WBI levels and WHO regions (Figure 1).

**Figure 1.**
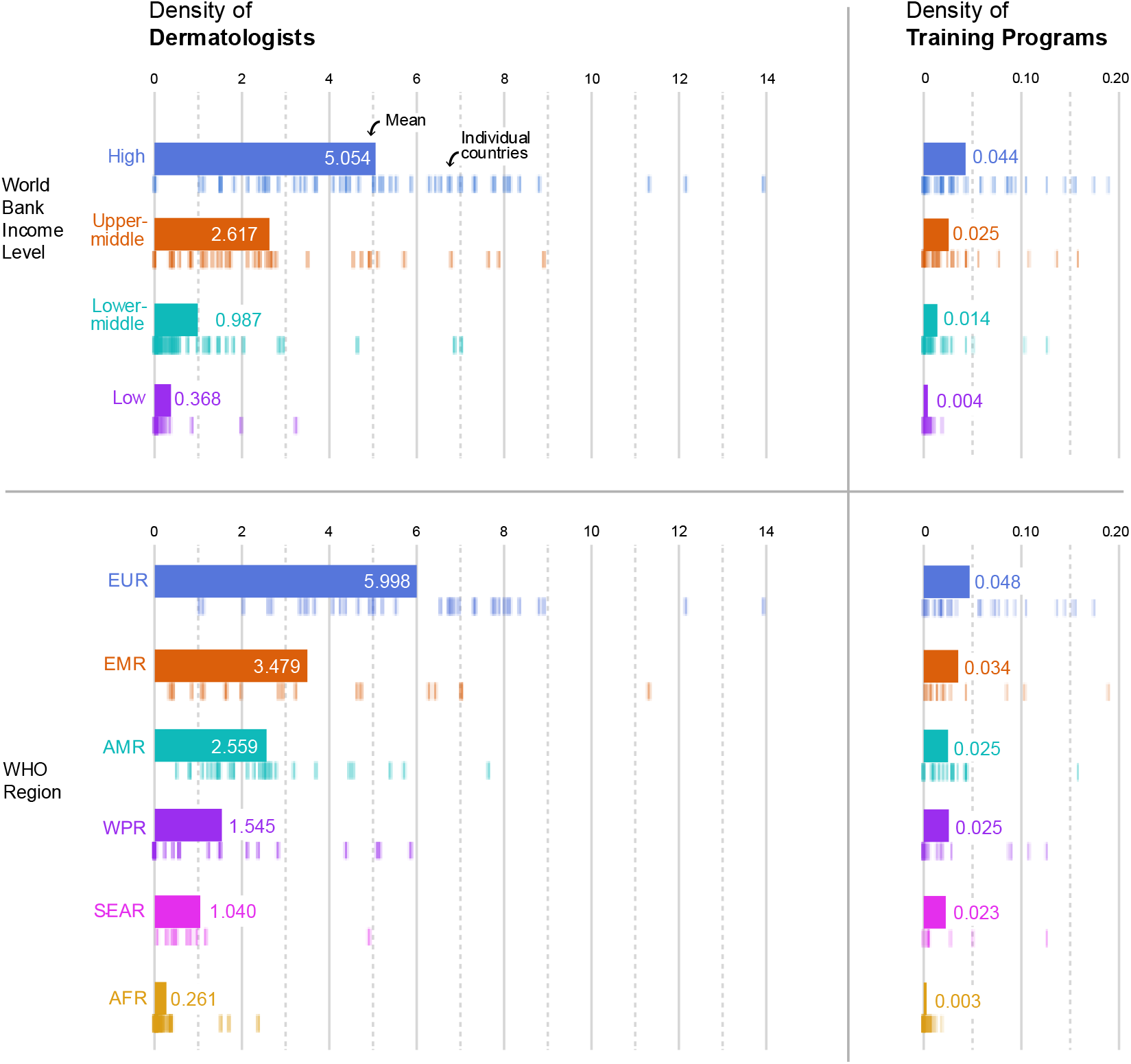
**Density of Dermatologists and Dermatology Training Programs, per 100,000 population**

Across the 158 responding countries, there were 169,366 reported dermatologists. Using the density of dermatologists across WHO regions and WBI levels in our sample, we estimate an unreported 6,267 dermatologists (95% prediction interval (PI): 4232-8302) in the missing 39 countries. This approximates a global population of 175,633 dermatologists (95% PI: 173,598-177,668).

Based on mean reported percentages, 79% of dermatologists worked in urban environments with significant variation (p < 0.001) across WBI levels (Figure 2).

**Figure 2.**
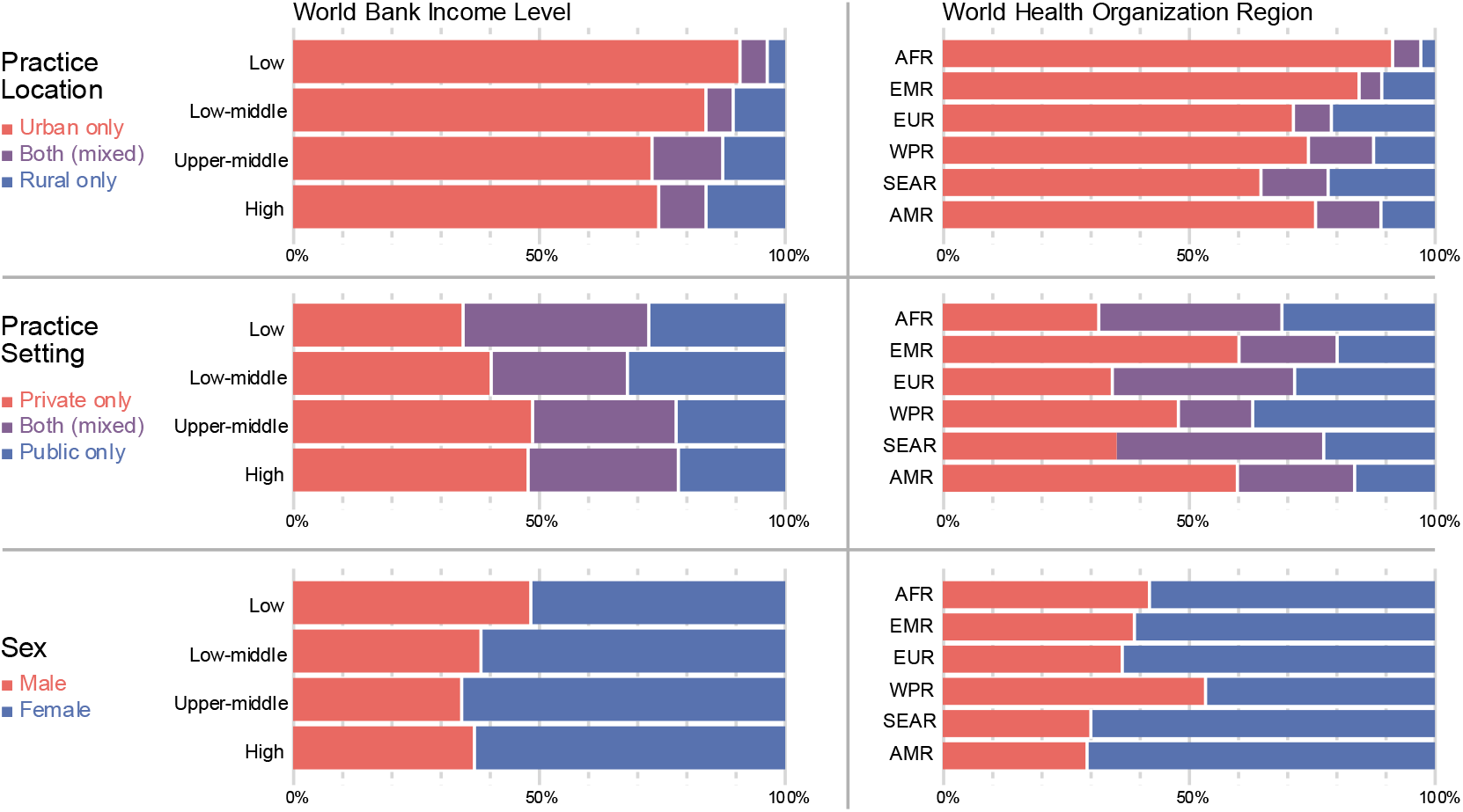
**Demographic Distribution of Dermatologists, by World Bank Income Level and World Health Organization Region**

Seventy-four percent of dermatologists in high-income countries and 73% in upper-middle-income countries worked in urban environments, versus 84% in lower-middle-income countries and 91% in low-income countries. For practice settings of dermatologists (Figure 2), a greater proportion of dermatologists in HICs practiced in strictly private settings (48% in high-income and 49% in upper-middle-income countries, versus 40% in lower-middle-income regions and 35% in low-income countries), although this was not statistically significant (p > 0.05). Lastly, 62% percent of reported dermatologists were female (Figure 2).

### Definition of a dermatologist and availability of training programmes

When evaluating how countries define a dermatologist, the majority (89%) required completion of both medical school and a formal dermatology residency, with little variation across WBI levels and WHO regions (Figure 3). Most countries also reported having a dermatologic governing body (82%). Most low- and lower-middle-income countries allow dermatologists to obtain training abroad (92%).

**Figure 3.**
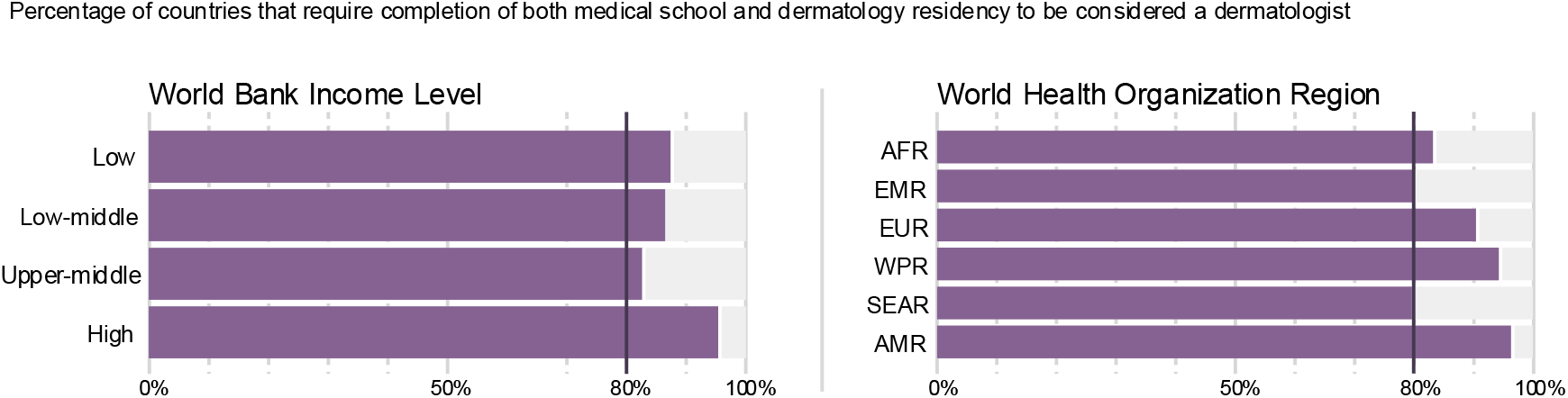
**Definition of a Dermatologist, by world Bank Income Level and World Health Organization Region**

Seventy-nine percent of countries reported having dermatology training programmes, with slight variation across WBI levels (78% in LICs, 73% in lower-middle, 78% in upper-middle, and 86% in upper-income countries). However, the density of training programmes varies significantly (p < 0.001) across WHO regions and WBI levels (Figure 1), with an average of 0.004 training programmes per 100,000 people in low-income countries, 0.014 in lower-middle, 0.025 in upper-middle, and 0.044 in high-income countries.

### Dermatology provider shortages, health infrastructure quality, and dermatological care access

Global shortages were noted for healthcare workers able to provide care for skin diseases, varying by WBI level (Supplement 2.3). The highest reported shortages were in LICs (89% of countries), while the lowest were in HICs (57%). There was similar variation in reported shortages across WHO regions, ranging from 94% of countries in AFR to 47% in EMR.

When rating healthcare infrastructure for adequate dermatological care, there was significant variation (p < 0.001) across WBI levels, with 61% of respondents in LICs reporting extremely poor or inadequate infrastructure versus 19% of HICs (Supplement 2.4).

When rating access to care, 41.6% of countries reported inadequate or extremely poor access to general dermatologic and 47.4% to specialist dermatologic care. Access to general, specialized, pediatric, surgical and dermatopathologic care varied significantly across WBI levels (p<0.001) (Figure 4). This general trend indicates that the vast majority of respondents in LICs reported insufficient access across all dermatology subtypes, with access to specialty care even more limited than general dermatological care.

**Figure 4.**
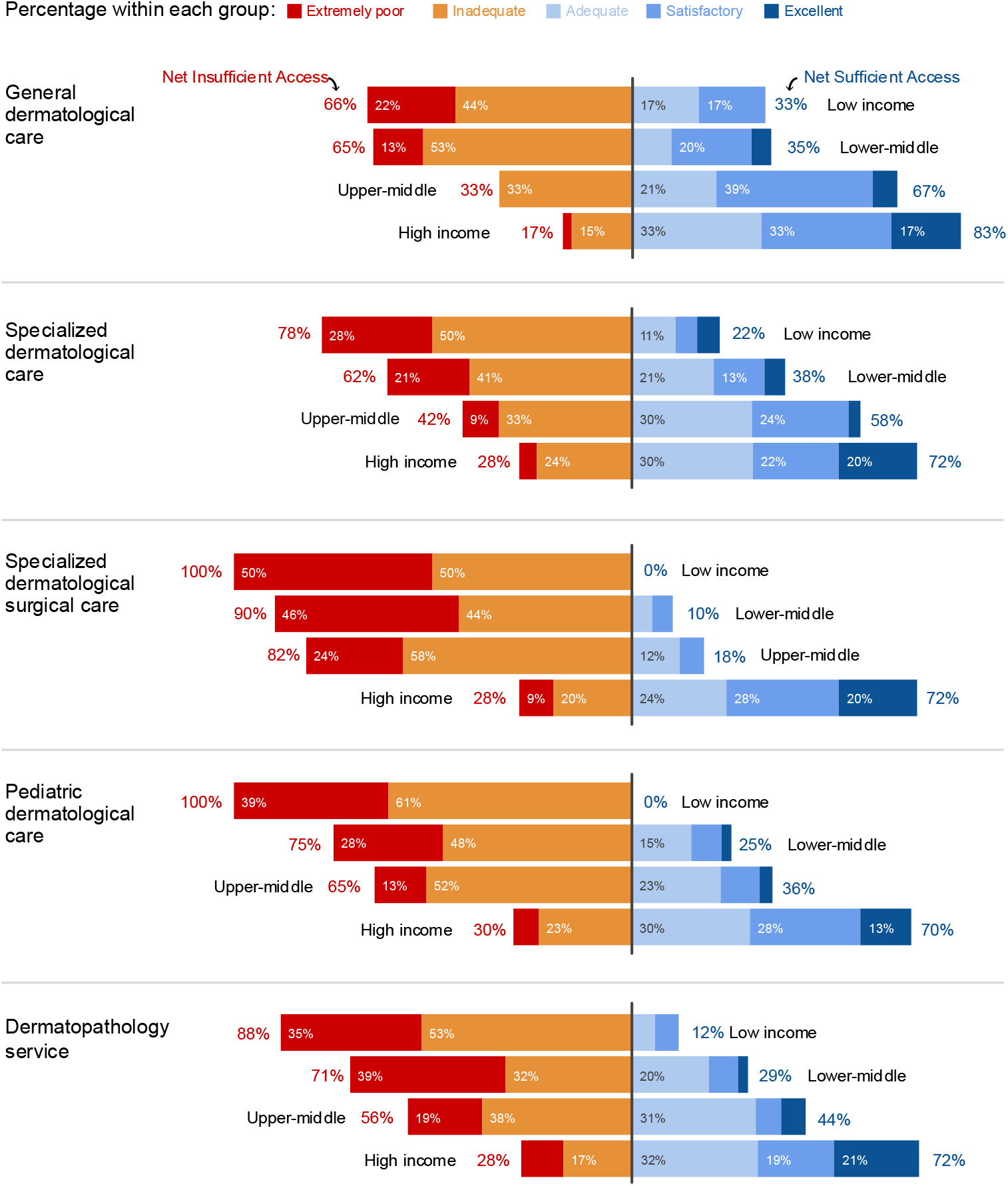
**How would you rate access to various types of dermatological care?**

### Care setting

Most dermatological care was delivered in an office-based setting (Supplements 2.5&2.6).

### Providers of dermatological care

Across all WBI levels, dermatologists and primary care physicians were most frequently reported as primarily responsible for diagnosing and treating skin diseases (Figure 5). Notably, in low- and lower-middle-income areas, a substantial amount of dermatological care is also delivered by health officers, pharmacists, nurses, traditional healers, and other medical specialists. When asked to rank the primary providers of care for skin diseases across WBI levels, dermatologists were consistently the most reported, followed by primary care physicians (Supplement 2.7).

**Figure 5.**
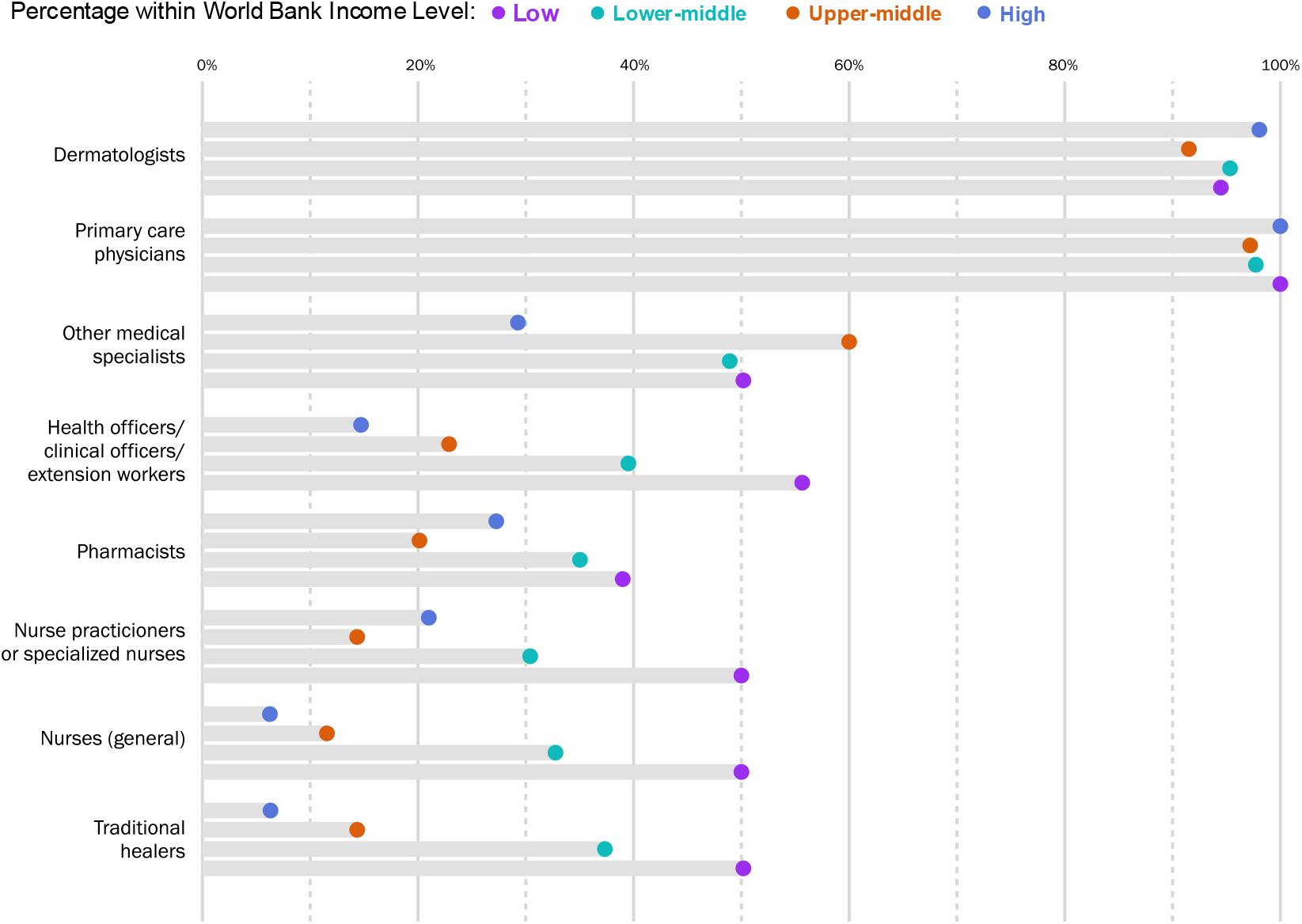
**Who is primarily responsible for the diagnosis and treatment of skin disease in your country?**

### Transition to cosmetics

Almost half of respondents (46%) reported access issues due to dermatologists transitioning to primarily cosmetic practices as a problem. There was significant variation (p = 0.006) across WBI levels, with only 17% of low and 35% of lower-middle-income countries reporting the transition to cosmetics as a problem, versus 56% of upper-middle- and 59% of HICs. Similar variation was found across WHO regions (Supplement 1.5).

### Government priority

Globally, a minority (26%) of responding countries reported that dermatological care is a national-level priority. Only 12% of LICs reported that skin disease is a government priority, compared to 50% in HICs (p = 0.152).

### Required number of dermatologists

Using an ‘adequate’ response to access to dermatological care (Figure 4) as a minimum threshold, 30 countries reported at least adequate access to all types of dermatological care. Using the mean density of dermatologists within this subpopulation, we estimate a minimum required density of 5.6 dermatologists per 100,000 people to serve a country’s population appropriately. Based on this density, HICs are 8.84 times (Odds Ratio (OR): 8.84, 95% CI: 3.42-22.83) more likely to have an adequate number of dermatologists to serve their population than countries in all other income categories.

Examining WHO regions with this same 5.6/100,000 density threshold, no countries in AFR or SEAR have enough dermatologists. In contrast, 51% of countries in EUR meet this threshold (Supplement 1.6).

## Discussion

We present a novel, systematic global assessment of access to dermatological care. Data from 158 countries, representing 96.7% of the global population, underscore considerable inequities in skin health. Forty-two percent of countries reported inadequate or extremely poor access to dermatological care and profound differences in dermatologist density across WBI levels.

### Trends in Access to Dermatological Care

Despite LMICs carrying a disproportionate burden of skin disease, two-thirds reported insufficient access to dermatological care, compared to only 17% of HICs.^2^ Historic structural inequities underlie access gaps in LMICs, resulting in training, workforce, and infrastructure deficits,^13, 31^ ultimately leading to significant delays in diagnosis and treatment, higher disease burden, and poor quality of life.^2, 32-34^

We found dermatologists cluster in urban areas (overall 79%), with misdistribution worsening as country income decreases. This mirrors previous studies documenting clustering in cities in U.S., Brazil, and Ethiopia.^10, 13^ In LMICs, we also found many dermatologists need to augment low public sector salaries with private practice. This ‘physician dual practice’^35^ may further reduce access as most patients receive care in public sectors.

HIC populations are not immune to ‘dermatological deserts,’ regions with limited access due to geographic clustering. More than half of HICs reported too few dermatologists, and less than one-third of HIC dermatologists work in public sectors. Prior HIC studies highlight substantive gaps in access to dermatological care for racial/ethnic minorities,^36^ people experiencing homelessness,^37^ and refugee/migrant populations.^38^ These gaps reinforce the concept of skin health access as multifaceted, requiring consideration of affordability (healthcare costs, insurance), accommodation (language, hours and locations), and acceptability (culturally sensitive care). Dermatologists transitioning to cosmetic-based practices exacerbate these inequities, with over half of respondents reporting this as a problem (59% in HICs).

### Dermatology Workforce

Overall, we estimate 175,633 dermatologists globally. The variability in density by country/region and WBI level is substantial (p<0.001), ranging from a mean of 0.37 per 100,000 people in LICs to 5.05 in HICs, with a mean density of 2.66 globally.

In working towards skin health equity, it is worthwhile to consider what constitutes a ‘sufficient’ dermatologist-to-population ratio. A 2001 survey study estimated a minimum of 3.3 dermatologists per 100,000 people. This estimate has considerable limitations, including reliance on a small, subjective U.S.-based sample which does not consider complexity of care delivery or global needs.^17, 39^

Based on countries reporting ‘sufficient’ access to general and specialty dermatological care, we present an updated estimate of at least 5.63 dermatologists per 100,000 to deliver adequate care. Only 14% of countries achieve this value, with 70% from EUR. Notably, countries above the threshold were more likely to have governments prioritizing dermatological care (p=0.007). HICs do not universally meet this bar. For example, the U.S. and Australia are below this value (4.47 and 2.38) with varied access to care for underserved populations.^17, 40^

The presence and retention of dermatologists are reliant upon in-country or regional training centers. Our data document dermatology residency training programmes as unevenly distributed by WBI level and WHO region, with 46% of AFR countries reporting no programmes. Dermatology training programmes are vital to expanding the workforce. Retaining trained clinicians is also critical as migration from LMICs to higher paying jobs in HICs amplifies workforce shortages.^41^

Investment in regional or in-country training programs that incentivize serving local populations can meaningfully improve access to care.^42^ The Regional Dermatology Training Centre in Tanzania^43^ – which has trained over 200 clinicians from 17 African countries – is an exemplar program.

Access to skin health relies on myriad factors from the patient level to international policy and requires global prioritization. In May 2025, the World Health Assembly passed (WHA7 .1) “Skin Disease as a Global Public Health Priority.” The resolution calls WHO member states to recognize the impact of skin disease and mandate national and international efforts to improve prevention, early detection, and equitable access.^1^ Our data document 74% of countries report skin health is not a governmental priority, underscoring the rigorous effort needed to achieve the resolution’s aims.

## Future Directions

### Advanced Training and Dermatological Subspecialty Care

Comprehensive dermatological care involves collaboration between subspecialties to provide expert-level histopathologic review, surgical interventions, and paediatric care. These needs are underscored by the largest increase in new cases of skin disease occurring in patients 0-4 years of age.^2^ Our data demonstrate substantial subspecialty care needs across WHO and WBI levels, particularly in LICs where 90% reported shortages in all dermatologic subspecialties.

### Methods of Alternative Care Delivery

Reflecting shortages of trained dermatologists and high disease burden, non-dermatologist clinicians consistently deliver dermatological care across all WHO regions and WBI levels. Primary care clinicians and paediatricians delivered specialized dermatological care in >50% of countries, with responsibility increasing as WBI level decreased.

Skin diseases are among the most common reasons for primary care and paediatrician visits.^45-47^ However, no standardized dermatological training for non-dermatologist healthcare workers exists, with many reporting only hours or days of education.^48, 49^ Limited training and profound case load put frontline healthcare workers at a disadvantage in diagnosing and treating skin disease.

In LMICs, health officers (44%), pharmacists (36%), specialized nurses (36%), general nurses (38%), and traditional healers (41%) also provide a substantial share of dermatological care. Despite their outsized role in low-resource settings, targeted formal dermatological training is even more limited.^50-53^

Task shifting expands dermatological access by reallocating specific diagnostic and management responsibilities to frontline providers.^52, 54, 55^ Successful models must be tailored to the specific roles, skillsets, and contexts of each cadre. For instance, in Uganda, skin biopsy training for Kaposi’s sarcoma diagnosis initially targeted primary care physicians but had greater success after including nurses and pharmacy technicians.^56^ Traditional healers can also be incorporated into disease recognition and referral protocols.^57-59^

Digital technologies and artificial intelligence (AI) can potentially improve access to care through teledermatology platforms, educational and decision-support tools, and online training. Teledermatology can overcome geographic/logistical barriers and reduce treatment delays via asynchronous and real-time platforms.^60^ However, availability and uptake are limited by infrastructure, disparities in technology access, regulatory challenges, and privacy concerns.^61, 62^

A growing ecosystem of digital educational and decision-support tools is emerging for non-dermatologist providers. Efforts including the WHO Skin NTDs mobile application and VisualDx provide evidence-based education and diagnostic resources for frontline providers.^53, 63^ The WHO Academy offers free digital educational content on multiple skin conditions.^64^ Other nascent AI-driven image recognition applications aim to triage lesions and detect high-risk findings,^65^ but are constrained by limited training data, poor performance in darker skin tones, and lack of transparency in development.^66, 67^

### Limitations

National dermatology leaders may have lacked access to verified statistics on their country’s workforce, with some respondents necessarily estimating numbers of dermatologists; to account for this, respondents indicated certainty levels and data sources for numerical values. The survey was limited to key informants and does not reflect patient experiences. The survey was only available in four languages, potentially limiting participation. Due to the survey’s broad reach, some questions may have been interpreted differently based on country context.

### Conclusion

Global access to dermatological care is a multifaceted health equity crisis shaped by national and international level prioritization, healthcare infrastructure, financial constraints and education and training. More trained dermatologists and other healthcare professionals equipped to treat skin diseases are needed across WBI levels and WHO regions, especially in LMICs and Africa, with incentives for rural and underserved practice. Interventions targeting frontline providers, paediatricians, nurses, traditional healers, and community workers, focused on common dermatoses, are a necessary and cost-effective way to improve access to quality dermatological care.

As the greatest burden – and often the most valuable insight – resides with patients and providers in resource-limited settings, these communities must be the starting point for meaningful solutions.

## Data Availability

A public data explorer will be available on www.skinobservatory.org. Additional data will be shared on reasonable request to the corresponding author.

## Acknowledgements

The authors would like to thank Jose Postigo from the World Health Organization for his valuable contributions to the design of this study. They would also like to thank Arpita Bhose, Lisa Jacobs, Amarni Wood, Emma Van Rooijen, and the rest of staff at the ILDS, as well as Nicole Holt, Ellie VanBlarcom, and Jodi Kurtz of the Medical Practice Evaluation Center at Massachusetts General Hospital for their assistance with the establishment of the SkinObservatory study. They would like to thank Tebyan Khalfalla, Parisa Shamaei Zadeh, Georgie McTigue, Natalie Asemi, and Morvarid Zehtab for contributions to data cleaning and data analysis.

