## Supplemental Materials for "Global Disparities in Access to Dermatological Care: the Skin Health Observatory"

<u>Contents</u>


1. **Additional Methods**
   1.1. WHO ‘Building Blocks’
   1.2. Addressing multiple country respondents and discrepancy adjudication
   1.3. Data sources - additional information
      1.3.1. Republic of Venezuela
      1.3.2. Chinese Taipei
      1.3.3. Niue and Cook Islands
   1.4. Additional statistical analyses methods – estimating the number of dermatologists worldwide and the corresponding prediction interval
   1.5. Transition to cosmetics
   1.6. Percentage of countries with an adequate number of dermatologists
2. **Additional Results**
   2.1. Supplemental Table 1 – Baseline demographics of survey respondents
   2.2. Supplemental Table 2 – Numbers of surveys submitted, by WHO region
   2.3. Supplemental Figure 1 - Proportion of countries reporting a shortage of dermatologists or dermatology subspecialists that can treat skin disease
   2.4. Supplemental Figure 2 - How would you rate country’s health infrastructure in providing dermatological care?
   2.5. Supplemental Figure 3 - In what setting are patients receiving dermatological care?
   2.6. Supplemental Figure 4 - In what setting are patients access specialized dermatological care?
   2.7. Supplemental Figure 5 – Who is primary responsible for diagnosing and treating skin disease in your country, ranked 1st, 2nd and 3rd?
3. **References**

### 1. Additional Methods

#### 1.1. WHO Building Blocks

The survey was developed to incorporate the WHO's health systems "Building Blocks" framework, which has been used to assess access to specialty care.[1-3] The framework offers a holistic approach for analyzing global access to care by arranging health systems into six components: leadership and governance; service delivery; health system financing; health workforce; health information systems; and access to essential medicines. Within each building block, key indicators are assessed.

#### 1.2. Addressing multiple country respondents and discrepancy adjudication

During survey results data cleaning, for countries with multiple respondents, discrepancies were resolved as follows: for numeric values, if the difference between answers was within 15%, the mean was taken. If the difference was >15%, countries were contacted for clarification. A similar approach was taken for ranking variables (from 1 to 5), but where estimates were close, mean was rounded to the most conservative estimate. Where in-country clarification for countries with multiple responses was sought but not able to be obtained, the answer where the respondent indicated the highest level of certainty was selected.

#### 1.3. Data Sources

##### 1.3.1. Republic of Venezuela

World Bank Income level classification was not available for Venezuela in 2024. Recent analyses by the Inter-American Development Bank have suggested that Venezuela is most likely a lower-middle income country – as such, this classification was adopted.[4]

##### 1.3.2. Chinese Taipei

Population estimates for Chinese Taipei were taken from the 2024 Chinese Directorate-General of Budget, Accounting and Statistics.[5]

##### 1.3.3. Niue and Cook Islands

The World Bank does not provide income level classification for Niue or the Cook Islands. As such, the WHO country overview income classification was used instead.[6,7]

#### 1.4. Additional statistical analyses methods – estimating the number of dermatologists worldwide and the corresponding prediction interval

To estimate the number of dermatologists worldwide, we predicted the number of dermatologists for countries that did not report a total using an interactive two-way ANOVA, with World Bank income level and WHO region as factors. We then summed these predicted values and added them to the total observed number of dermatologists. To calculate this estimate’s 95% prediction interval, we summed the predicted variances and applied the finite population correction (FPC) factor and normal distribution critical values.

#### 1.5. Transition to cosmetics

There was significant variation in accessing dermatological care due to too many dermatologists transitioning to primarily cosmetic practices ($p < 0.000$) across WHO regions. There was less of a reported burden in the African (6% of countries), South-East Asian (40%), and Western Pacific (41%) regions versus the European (61% of countries), Americas (68%), and Eastern Mediterranean (73%) countries. Among respondents who ranked the problem's severity (36 countries), 78% ranked it as at least a moderate problem.

#### 1.6. Percentage of countries with an adequate number of dermatologists

Using 5.6 dermatologists per 100,000 people as a threshold to provide adequate access to dermatologic care across WHO regions 0% of countries in the African region, 0% of countries in the South-East Asian region, 5% of countries in the Western Pacific region, 7% of countries in the Americas, 26% of countries in the Eastern Mediterranean, and 51% of countries in the European region meet this threshold.

### 2. Additional Results

#### 2.1. Supplementary Table 1

**Demographics of survey respondents**

| | | |
|---|---|---|
| Total number of surveys submitted | | 195 |
| Age group (%) | Under 20 | 0 |
| | 20-29 | 0 |
| | 30-39 | 23 (12.6) |
| | 40-49 | 65 (35.5) |
| | 50-59 | 55 (30.1) |
| | 60-69 | 34 (18.6) |
| | 70 and over | 6 (3.3) |
| | Missing | 12 |
| Sex (%) | Male | 98(52.1) |
| | Female | 88(46.8) |
| | Other/prefer not to say | 2 (1.1) |
| | Missing | 12 |
| Role (%) | Dermatologist (including dermatologist presidents of national societies) | 172 (91.5) |
| | Dermatology student/trainee | 3 (1.6) |
| | National program director/Administrative president of nation dermatology society | 3 (1.6) |
| | Dermatovenereology officer | 3 (1.6) |
| | Physician (non-dermatologist) | 4 (2.1) |
| | WHO technical officer | 1 (0.5) |
| | Public health officer/ researcher | 2 (1.1) |
| | Missing | 12 |

2.2. Supplemental Table 2

**Numbers of surveys submitted by each country**

| | | Number of surveys submitted | | |
|---|---|---|---|---|
| | | 1 | 2 | 3 or more |
| Total number of countries | | 129 | 23 | 6 |
| WHO regions | AFR | 31 (Angola, Burundi, Burkina Faso, Central African Republic, Chad, Côte d'Ivoire, Cameroon, Republic of Congo, Cabo Verde, Algeria, Eswatini, Ethiopia, Gabon, Ghana, Guinea, The Gambia, Liberia, Lesotho, Madagascar, Mali, Mozambique, Mauritania, Mauritius, Namibia, Niger, Nigeria, Senegal, Sierra Leone, Tanzania, Uganda, Zimbabwe) | 5 (Benin, Kenya, South Africa, Togo, Zambia) | 4 (Botswana, Democratic Republic of Congo, Malawi, Rwanda) |
| | EMR | 12 (Afghanistan, Arab Republic of Egypt, Djibouti, Iraq, Islamic Republic of Iran, Kuwait, Libya, Morocco, Republic of Yemen, Saudi Arabia, Syrian Arab Republic, Tunisia) | 7 (Bahrain, Jordan, Lebanon, Occupied Palestinian Territory, Pakistan, Sudan, United Arab Emirates) | 0 |
| | EUR | 32 (Andorra, Austria, Belgium, Belarus, Croatia, Cyprus, Czechia, Germany, Denmark, Estonia, Finland, Georgia, Hungary, Ireland, Italy, Kazakhstan, Malta, Netherlands, North Macedonia, Norway, Poland, Portugal, Romania, Russian Federation, Slovak Republic, Spain, Sweden, Tajikistan, Türkiye, United Kingdom, Uzbekistan) | 4 (Switzerland, Greece, Israel, Serbia) | 1 (France) |
| | WPR | 19 (Cambodia, China, Chinese Taipei, Fiji, Hong Kong Special Administrative Region of the People's Republic of China, Japan, Kiribati, Marshall Islands, | 2 (Australia, Solomon Islands) | 1 (Malaysia) |

| | | | |
|---|---|---|---|
| | Mongolia, Naura, New Zealand, Philippines, Palau, Papua New Guinea, Samoa, Singapore, Tonga, Republic of Korea, Vanuatu) | | |
| SEAR | 9 (Bangladesh, Bhutan, India, Maldives, Myanmar, Nepal, Sri Lanka, Thailand, Timor-Leste) | 1 (Indonesia) | 0 |
| AMR | 26 (Argentina, Antigua and Barbuda, Belize, Bolivia, Bolivian Republic of Venezuela, Brazil, Barbados, Canada, Chile, Colombia, Costa Rica, Cuba, Dominican Republic, El Salvador, Honduras, Haiti, St. Kitts and Nevis, St. Lucia, Mexico, Nicaragua, Panama, Peru, Suriname, The Bahamas, Uruguay, United States) | 4 (Ecuador, Guatemala, Jamaica, Paraguay) | 0 |

AFR = Africa Region, AMR = Americas Region, EMR = Eastern Mediterranean Region, EUR= European Region, SEAR = South-East Asia Region, WPR = Western Pacific Region

### 2.3 Supplemental Figure 1

**Proportion of countries reporting a shortage of dermatologists or dermatology subspecialists that can treat skin disease.**

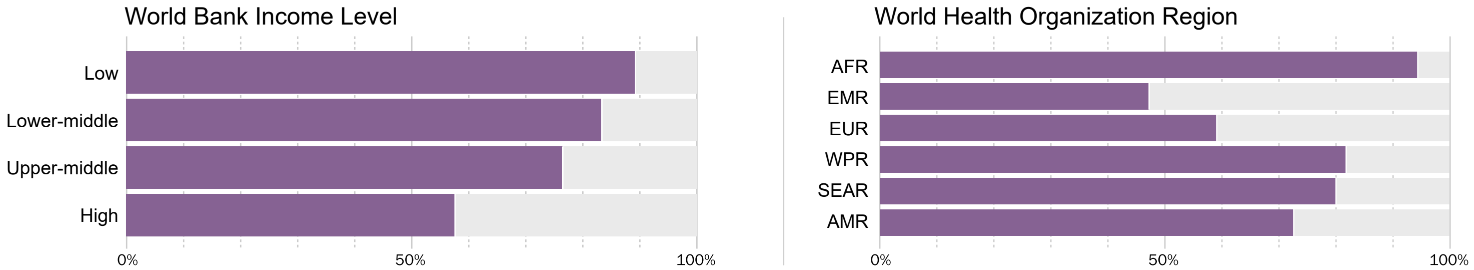

### 2.4 Supplemental Figure 2

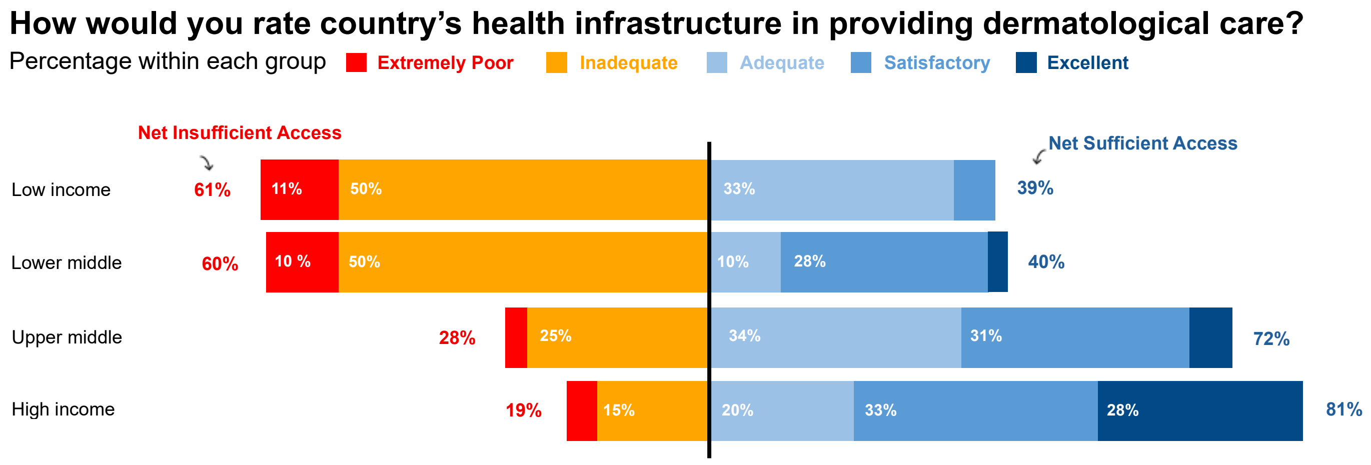

### 2.5 Supplemental Figure 3

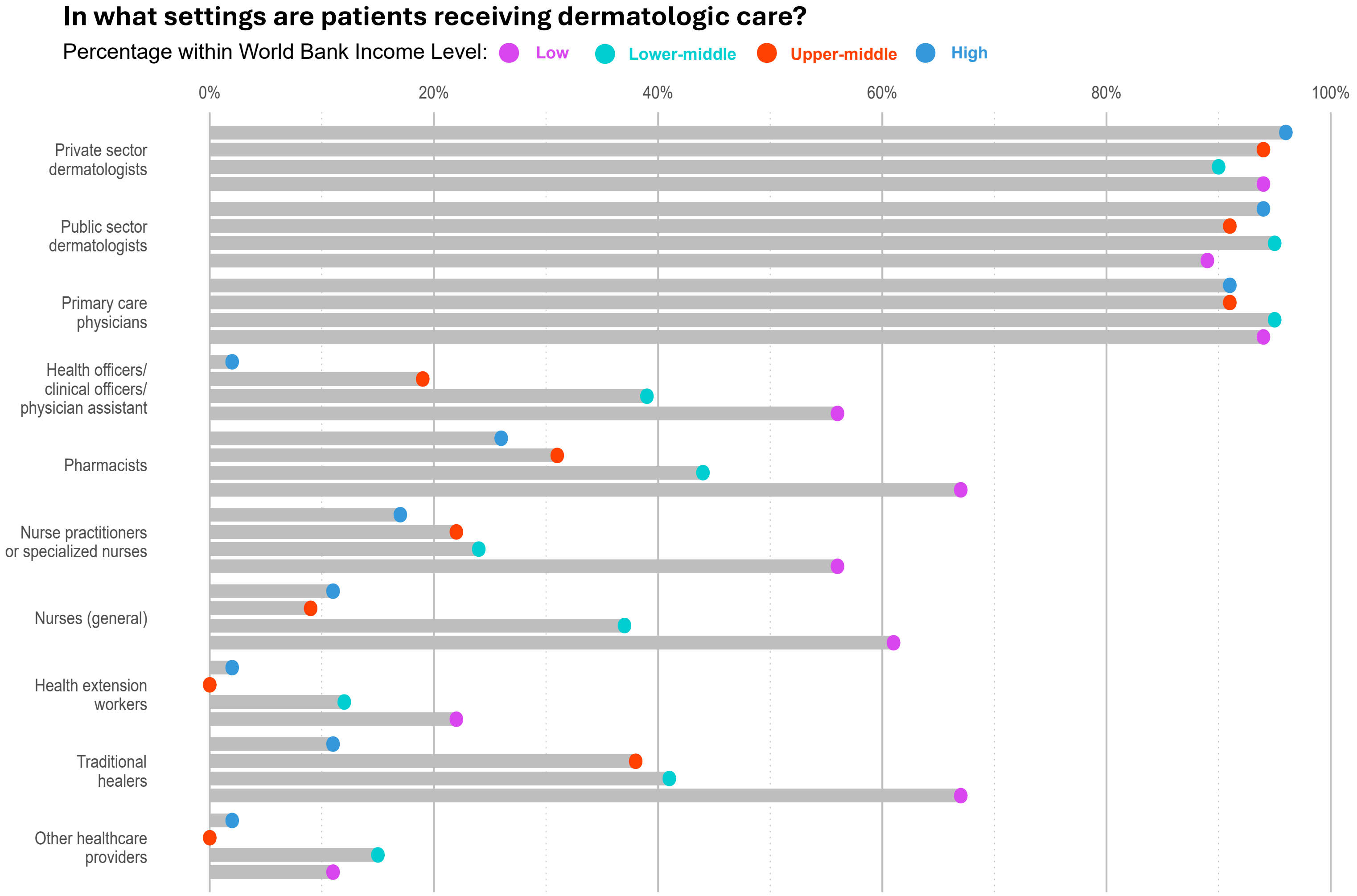

### 2.6 Supplemental Figure 4

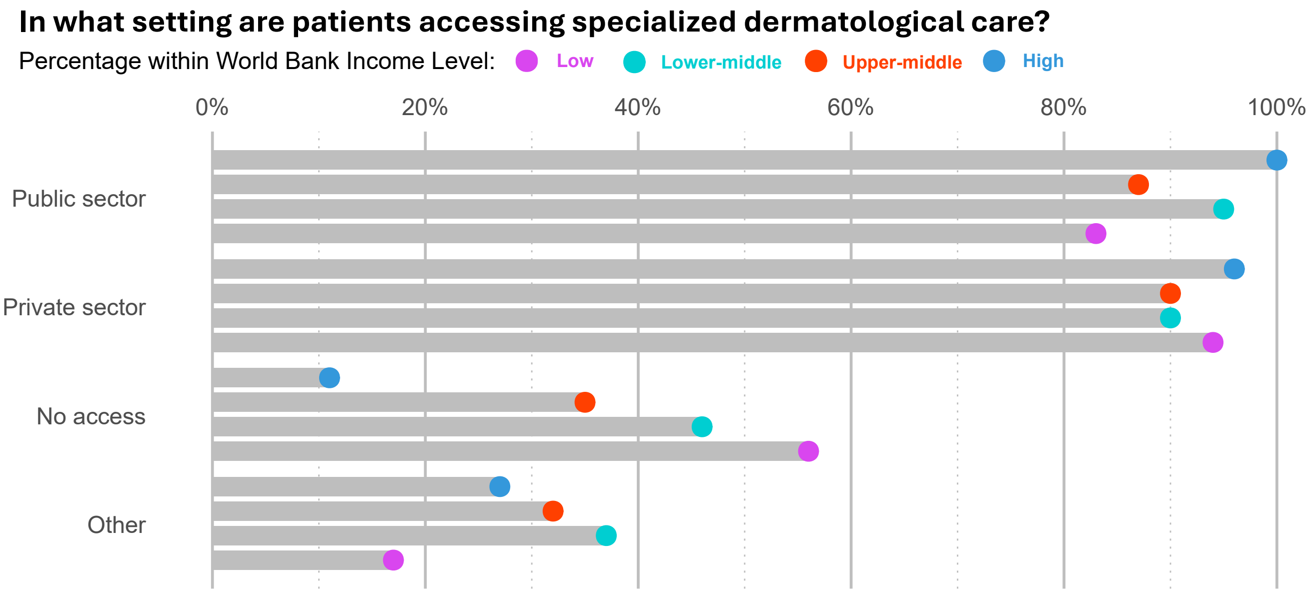

### 2.7 Supplemental Figure 5

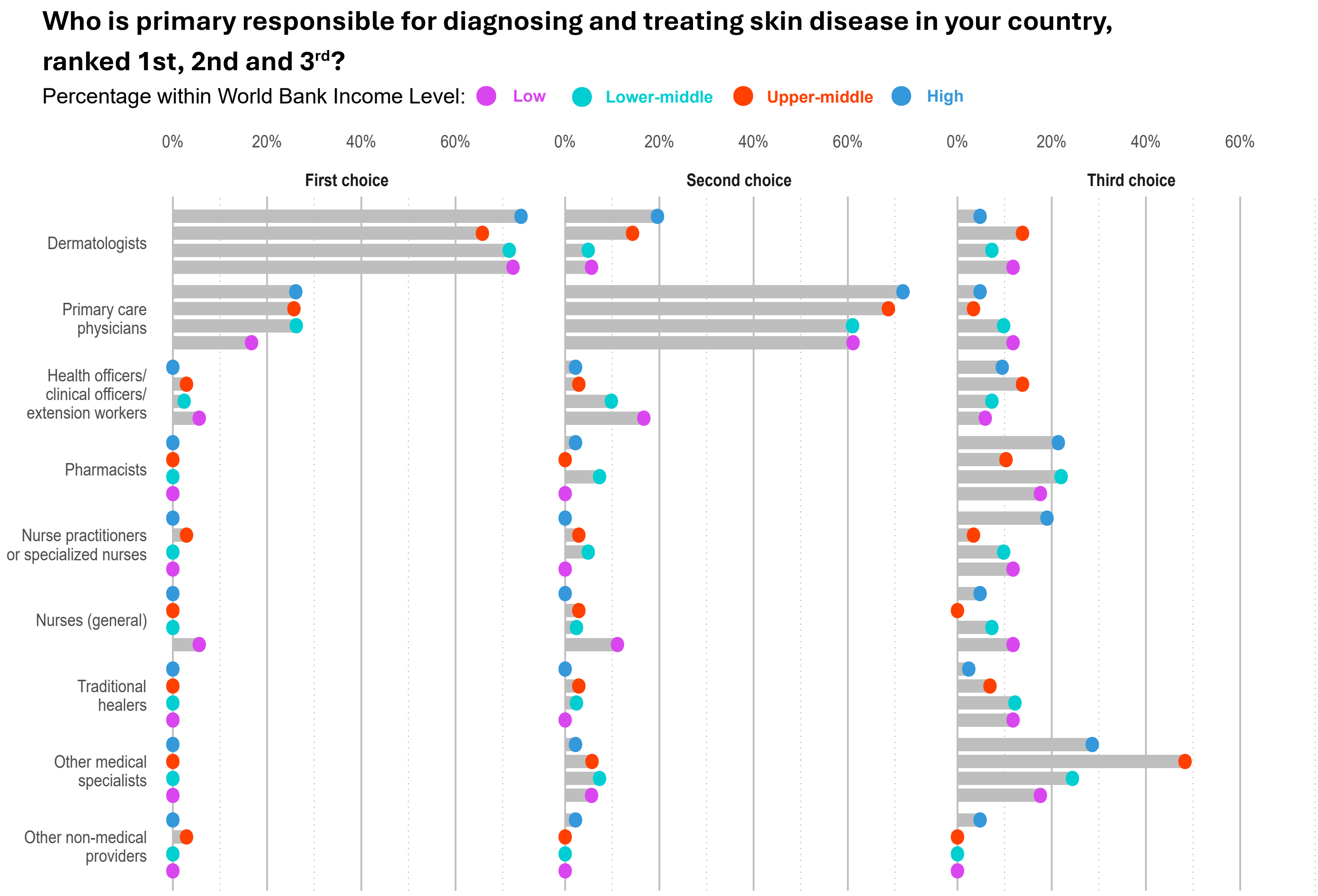
